# Effects of a 12-week unsupervised and adapted physical activity prescription on physical performance in geriatric outpatients with overweight or obesity: the PACE tool

**DOI:** 10.64898/2026.09.06.26359363

**Authors:** H El-Oueslati, L Youssef, Ruiz F-J-M, L Boucher, T Tannou, C Brodeur, F Andriamampionona, Kergoat M-J, M Aubertin-Leheudre

**Affiliations:** Département des sciences de l’activité physique, Groupe de recherche en activité physique adapté, Université du Québec à Montréal, Montréal, Québec-Canada; Centre de Recherche de l’Institut Universitaire de Gériatrie de Montréal, CIUSSS du Centre-Sud de l’Île-de-Montréal, Montréal, Québec- Canada; Département de Médecine, Faculté de médecine, Université de Montréal, Montréal, Québec- Canada; École de Kinésiologie et des Sciences de l’Activité Physique, Faculté de médecine, Université de Montréal, Montréal, Québec- Canada

**Keywords:** exercise, sarcopenia, frailty, fat-mass, aging

## Abstract

**Background:** Sarcopenia and obesity often coexist in older adults and are associated with higher functional decline, disability, and reduced quality of life. Although physical activity (PA) is protective, prescription and adherence to exercise programs remains low in this population. Pragmatic and scalable interventions are therefore needed. Thus, we evaluated the effects of a 12-week unsupervised PA prescription program on physical performance in overweight/obese frail geriatric outpatients.

**Methods:** A single-arm interventional study was conducted among geriatric outpatients aged ≥60 years. Thirty-seven overweight/obese participants (20 women) completed the 12-week home-based PACE prescription which included individualized functional and resistance exercises with walking. Physical performance [i.e. functional capacity (SPPB), walking speed, Timed Up and Go (TUG), lower-limb muscle endurance (30-STS) and function (5-STS)], clinical status (probable sarcopenia, fall risk, walking speed) and clinically meaningful changes were assessed.

**Results:** Participants achieved 238 ± 140 min/week of PA on average, with 65% meeting recommended PA guidelines. Significant improvements were observed in lower-limb muscle function (*p*=0.033) and endurance (*p*=0.006) and in absolute (*p*=0.038) or relative (*p*=0.045) estimated muscle power. The prevalence of probable sarcopenia decreased significantly following the intervention (*p*=0.011). Clinically meaningful improvements were observed in muscle endurance (*p*<0.001) and functional status (*p*=0.040).

**Conclusion:** In overweight/obese frail geriatric outpatients, the prescription of PACE achieved high PA adherence and improved lower-limb muscle function. Integrating this pragmatic and personalized PA strategy into routine geriatric care may support healthy aging and help prevent sarcopenia-obesity related functional decline.

**Key points:**

- A 12-week unsupervised and adapted PA prescription (PACE tool) is feasible in frail geriatric outpatients with overweight or obesity.
- High adherence was achieved, with 65% of participants meeting or exceeding PA recommendations, alongside significant improvements observed in lower-limb muscle function (power and endurance).
- The intervention led to a clinically meaningful reduction in probable sarcopenia prevalence.
- PACE could represent a pragmatic and scalable strategy to improve functional outcomes in overweight/obese geriatric population.

## INTRODUCTION

Sarcopenia and frailty are prevalent age-related conditions leading to physical declines. Sarcopenia is characterized by the loss of skeletal muscle mass and function, whereas frailty is a clinical syndrome marked by reduced physiological reserves and increased vulnerability to stressors ^1^. Both conditions share core features, including low muscle mass, low grip strength, and slow gait speed, which together heighten susceptibility to adverse outcomes in older adults ^1^. Notably, impaired muscle function is strongly linked to impaired physical performance, a loss of independence in daily activities ^2^, and a higher all-cause mortality ^3^.

Alongside muscle deterioration, aging is often accompanied by increased adiposity. The coexistence of low muscle mass and excess fat accumulation has led to the recognition of sarcopenic obesity, now regarded as a distinct clinical entity. Beyond simple coexistence, adiposity may exacerbate muscle degeneration through metabolic alterations ^4^. Visceral fat accumulation is known to elevate levels of pro-inflammatory markers which contribute to accelerate muscle loss ^5,6^ as well as intramyocellular lipid deposition which could compromise skeletal muscle metabolic function and thereby limiting its hypertrophic capacity ^7–9^. Consistent with these mechanisms, obesity-related intramuscular fat infiltration has been associated with muscle weakness and accelerated functional decline ^10,11^. Being obese is also associated with an increased risk of falls in older adults, as well as a higher likelihood of disability in activities of daily living following a fall ^12^. Consequently, the coexistence of obesity and muscle dysfunction may accelerate physical deterioration, elevate the risk of hospitalization, and contribute to earlier loss of independence in older adults ^4,13^.

Physical inactivity is another major contributor of age- and obesity-related impairments ^14^. The World Health Organization recognizes that insufficient physical activity (PA) is a key modifiable risk factor for non-communicable diseases ^15^. In older adults, sedentary behavior contributes to impaired skeletal muscle metabolism, fat accumulation and progressive loss of muscle function ^16,17^. However, low PA levels and high sedentary behavior are prevalent in older adults ^16^. Moreover, epidemiological data indicate that up to 57% of adults with overweight or obesity report no weekly PA, and nearly 90% fail to achieve the minimum recommended 150 minutes per week ^18^. These findings highlight a significant gap between public health recommendations and actual behavior. Multiple factors contribute to this gap. Beyond biomechanical limitations and reduced exercise tolerance, individuals with obesity often experience several barriers such as internalized weight stigma, teasing and discrimination, which might discourage their engagement in group-based or supervised exercise settings ^19,20^.

Despite the strong evidences that support the benefits of regular PA in attenuating aging-related multi-system deterioration ^17,21^, integrating PA into the usual care of older outpatients with health conditions remains challenging. The main older adults barriers are the exercise intervention modality (supervised), physical or financial limitations, limited facility availability, adverse weather, time constraints, or the intimidating gym environment ^22^. Furthermore, prescribing PA within healthcare is uncommon due to the lack of pragmatic implementation strategies ^23^. In this context, unsupervised tailored and adapted exercise interventions offer a practical and potentially effective alternative ^24^, highlighting the need for scalable approaches that enable individualized PA prescriptions without requiring supervision, specialized equipment, or transportation for the patient and specific knowledge or burden of time for the physician.

To address this gap, the PACE tool (Promote the Autonomy through exerCisE) was co-designed as a structured and adapted PA prescription in outpatient settings. The tool integrates validated subjective and objective decisional trees (ODT) assessing mobility profile and allowing tailored programs and clinical implementation ^25,26^. Therefore, the objectives of this study were: (i) to assess PA adherence among frail geriatric outpatients with overweight or obesity and (ii) to evaluate the impact of a 12-week PACE-based PA prescription on physical performance and on their clinical changes.

## METHODS

### Study design

This *a-posteriori* per-protocol analysis was conducted using data from a single-arm interventional study conducted from April 2022 to April 2024 in the geriatric outpatient clinics of the Institut Universitaire de Gériatrie de Montréal (IUGM). The study protocol was approved by the IUGM Ethics Committee (#CER-VN-20-21-06), and all eligible participants provided verbal informed consent prior to enrollment in the PA program.

### Participants

Among 103 eligible geriatric outpatients (>60 years old) initially assessed at baseline (T0), 57 (55%) were classified with overweight or obesity (body mass index > 25 kg/m²). Of the 57 eligible participants, 13 (22%) withdrew during the 12-week unsupervised home-based intervention. The main reasons are unavailability or travel (n=11), knee joint pain (n=1), and liver-related health issues (n=1). Consequently, 44 overweight-obese participants followed the PACE program and underwent pre- and post-(T12) intervention assessments. During data processing, five participants had missing data for key outcomes and two participants were identified as outliers. Therefore, the final per-protocol analysis included 37 participants (women: n=20; men: n=17; see **Supplemental Figure 1**).

Initial inclusion criteria were as follows: (a) age ≥ 60 years; (b) absence of contraindications to performing PA in either sitting or standing positions; (c) ability to communicate in French or English; (d) ability to attend the clinic in person or to have access to videoconferencing technologies (e.g., Zoom©); (e) no severe cognitive impairment (i.e. able to collaborate); and (f) not receiving any rehabilitation services.

### PACE tool and PACE intervention overview

The pragmatic PACE tool includes 35 individualized, unsupervised, and adapted prescribed PA programs integrating multimodal exercises (min-max: 2-4) targeting strength, balance, and stability, alongside specific daily walking which was developed for outpatient geriatric clinics following a creation methodology.

Briefly, during usual geriatric consultations, the physician administered a subjective decisional tree (SDT) (13 questions derivate from FiND, FRAIL and SARC-F tool; score: x/18) to generate a 12-week, home-based, unsupervised and adapted PA prescription from the mobility profile score obtained. To ensure safety and appropriateness of each prescribed program, a kinesiologist (exercise certified trainer) administered an ODT [score: x/18; 4 validated tests: 1) 30-sec sit-to-stand; 2) bipodal balance; 3) Functional Reach Test; and 4) 4-meter walking speed] and teached the program remotely or in person. Compared to the pilot phase ^25^ and according to pragmatic co-created approaches (based on feedback and findings), three modalities (easier, recommended and advanced option) per exercise are now included in this revised version to ensure adherence and both decisional tree scores were expanded from 15 to 18 as maximum score to better identify the mobility profile as shown in ^26^.

Moreover, during the 12-week PACE intervention, participants completed a logbook including the number of exercises performed, perceived level of difficulty (on a 4-point Likert scale) and walking duration. Phone calls were also conducted at weeks 1, 3, 6, 9 to collect data on exercise habits details from and outside the prescription. At week 12, the ODT and exercise follow-up were assessed both in person and remotely (see **Supplemental Figure 2** for more details regarding the PACE intervention).

### Measurements

#### 1) Feasibility and acceptability

- *<u>Feasibility:</u>* was assessed through adherence to the PACE program. Weekly exercise volume and weekly walking volume were recorded and summed to calculate total weekly PA volume, expressed in minutes per week (min/week).
- *Acceptability:* was evaluated at the end of PACE intervention using the system usability scale (SUS) questionnaire, with a score ≥ 70/100 considered acceptable ^27^.

#### 2) Physical performance

More details about the following assessments were previously described in *Peyrusqué et al.* ^28^.

- *<u>Functional capacities (SPPB: x/12):</u>* were assessed using the validated Short Physical Performance Battery (SPPB), which includes 3 sub-tests (bipodal balance test, usual 4-meter walking test and 5-repetition sit-to-stand test (5-STS). The total SPPB score ranges from 0 (severe disability) to 12 (no disability), with a 1-point change considered as a minimum clinically important difference (MCID).
- *Normal walking speed (m/sec)* was evaluated using the validated 4-meter walk test, where a difference of 0.1 m/s is considered as MCID.
- *Gait parameter (sec):* was assessed using the usual pace 3-meter Timed Up & Go (3m-TUG) test where a time >14 seconds indicates a fall risk.
- *<u>Lower-limb muscle function:</u>* was assessed using the 5-STS. Performance was quantified as the time required to complete the test (s).
*<u>Lower-limb muscle endurance</u>:* was assessed using the 30-second sit-to-stand test (30-STS). Performance was expressed as the total number of repetitions completed, with a 1-point change considered as MCID.
- *<u>Lower-limb muscle power:</u>* was estimated using the below and validated STS power equation (see ^29^ ^for^ ^more^ ^dertails)^. Lower-limb muscle power was expressed in absolute (W) and in relative (W/kg). Relative power improvements exceeding the minimal clinically important difference thresholds of 0.33 W·kg⁻¹ for women and 0.42 W·kg⁻¹ for men were considered as MCIDs ^29^.

#### 3) Clinical categorization & clinically meaningful improvements

To facilitate the clinical interpretation of intervention-related changes, several categorical variables were derived from continuous outcome measures using established clinical cut-off values.

- *<u>Probable sarcopenia</u> (sarcopenic vs. non-sarcopenic)*: identified according to the EWGSOP2 criteria, whereby participants required ≥15 s to complete the 5-STS.
- *<u>Fall risk</u> (fall risk vs. not at risk)*: defined as a TUG performance ≥14 s.
- *<u>Slow walking speed</u> (slow walking vs. normal walking)*: slow walking speed was identified for participants presenting a usual walking speed <0.8 m·s⁻¹.
- *Lower limb muscle endurance:* Participants demonstrating an increase of ≥2 repetitions on the 30-STS were classified as achieving a clinical improvement.
- *Functional performance:* Participants exhibiting an improvement of ≥1 point on SPPB were classified as achieving a clinical improvement.

### Statistical analysis

Continuous variables are presented as mean ± standard deviation (SD), whereas nominal variables are reported as counts and percentages. Descriptive statistics were initially reported as counts and percentages for clinical categorial outcomes. Wilcoxon signed-rank test was used to evaluate the effect of the PACE program on physical performance outcomes pre- (T0) and post-intervention (T12). Wilcoxon effect size was given by the matched rank biserial correlation and can be interpreted as follows: small (0.1–0.3), moderate (0.3– 0.5) or large (>0.5). Changes in categorical clinical status between pre- and post-intervention were assessed using non-parametric McNemar’s test for paired nominal data to determine whether the proportion of participants in each category differed significantly between time points for probable sarcopenia status, fall risk and walking speed statutes. To determine meaningful clinical change for muscle endurance and functional performance statutes following the intervention, variables were categorized as improvement, unchanged, or deterioration based on predefined validated thresholds and evaluated using an exact binomial test. In our analysis, both improvement or unchanged statutes were considered as a favorable change given that, in geriatric populations, maintaining and preventing decline are considered clinically relevant to preserve aging trajectory ^30^. All statistical analyses were generated using JASP software (Version 0.95.4.0; JASP Team, 2019). Statistical significance was set at *p* < 0.05.

## RESULTS

Baseline characteristics including age, BMI, functional autonomy (ADL/IADL), cognition, and mental health were similar between participants who completed the intervention and those who drop-outs (**Supplemental Table 1**).

### Feasibility and acceptability of PACE program

Weekly engagement in the PACE program averaged 72 ± 59 minutes [range (min-max): 0-320 min/week] for exercise and 171 ± 125 minutes [range (min-max): 19-554 min/week] for walking, resulting in a total weekly PA volume of 244 ± 140 min/week [range (min-max): 51-577 min/week]. Overall, nearly two-thirds of participants (65%) accumulated more than 150 minutes of moderate physical activity per week.

The program prescribed is based on ODT dissociating impairment focus (color: n= 7) and difficulty level (number from I to V) to target a perceived intensity as “moderate”. Thus, among the PACE program prescribed, 41% of the participants (n=15) received the blue program which focussed on muscular function (e.g., sit-to-stands, heel-to-toe raises, squats, knee and arm lifts, static leg pushes), 16% and 13% received the yellow and the red which targeted the balance and the flexibility respectively. Finally, 16% followed the brown program which aimed to work each domain of the blue, the red and the yellow programs. No orange program was prescribed. Regarding the intensity, all the levels were prescribed but mainly level III (n=13/37: 35%) and V (n=13/37: 35%). Finally, the average daily walking time prescribed was 25 ± 6 minutes with the majority of participants (n=22/37: 59%) having 30 min of daily walk as prescription (see **Supplemental Figure 3** for more details).

The PACE intervention achieved a mean SUS score of 75/100, exceeding the commonly accepted usability threshold. Moreover, more than two-thirds of participants (68%) perceived the intervention as acceptable according to established SUS interpretative guidelines (score > 70/100).

### Statistical changes following PACE intervention

The effects of PACE program on physical performance are summarized in **Table 1 and Figure 1**. Lower-limb muscle function demonstrated statistical changes (5-STS; pre: 16.1 ± 5.4 vs. post: 14.9 ± 5.9, *p* = 0.033; 30-STS; pre: 9.9 ± 3.2 vs. post: 11.1 ± 3.8, *p* = 0.006) following the intervention. These improvements were accompanied by significant increases in both absolute (pre: 166 ± 76 vs. post: 181 ± 84, *p* = 0.038) and relative STS-derived muscle power (pre: 2.15 ± 0.84 vs. post: 2.34 ± 0.97, *p* = 0.045). However, functional capacity (SPPB total-score) and walking speed didn’t change significantly (all *ps* > 0.05), as confirmed by the small ESs (−0.22 and 0.17, respectively). Although gait parameter (TUG) performance did not reach statistical significance (but tend; *p* = 0.06), a moderate ES (0.37) suggests a potential reduction in fall risk following the intervention.

**Figure 1.**
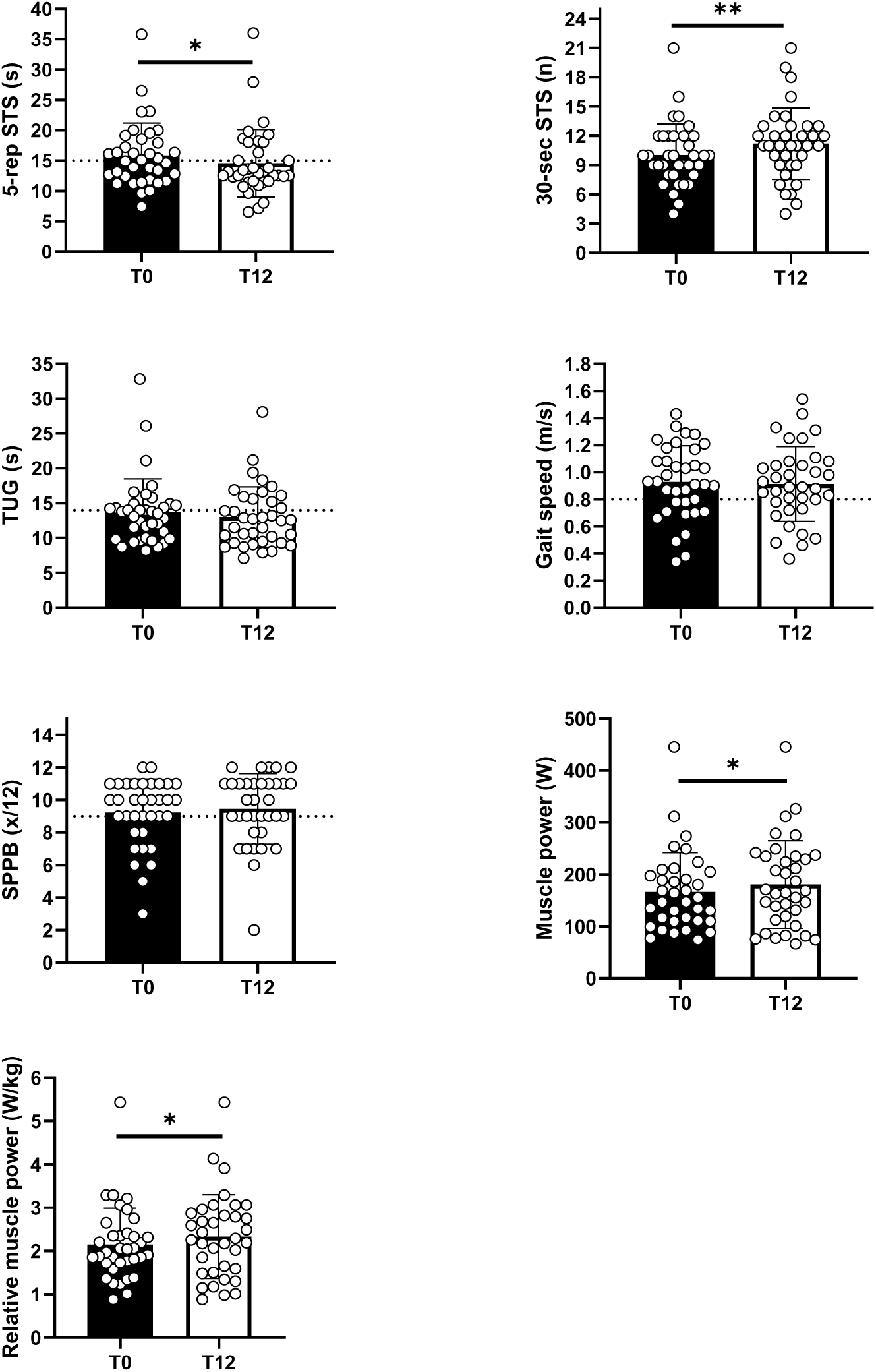
Effect of a 12-weeks PACE program on physical performance outcomes in frail obese geriatric outpatients. Results are presented as mean ± SD. T0 = baseline assessment. T12 = 12-week assessment. SPPB = short physical performance battery; TUG = timed up and go; 30-sec STS = 30 seconds sit-to-stand test; 5-rep STS = 5 repetitions sit-to-stand test; *: *p* < 0.05; **: *p* < 0.01. Black bar refers to baseline assessment (T0) and white bar refers to the 12-week assessment (T12).

**Table 1.** Physical performance adaptations following the 12-week PACE intervention in frail obese geriatric outpatients.

| Outcomes | n | Pre | Post | *P*-value | ES | Change (Δ) | Level of change |
| --- | --- | --- | --- | --- | --- | --- | --- |
| Muscle strength (5-STs; s) | 36 | 16.1 ± 5.4 | 14.9 ± 5.9 | **0.033\*** | 0.42 | -1.18 | + |
| Muscle endurance (30-STs; n) | 36 | 9.9 ± 3.2 | 11.1 ± 3.8 | **0.006\*\*** | -0.61 | +1.17 | + |
| Muscle power (MP; W) | 36 | 166 ± 76 | 181 ± 84 | **0.038\*** | -0.46 | +14.51 | NA |
| Relative MP: All (W/kg) | 36 | 2.15 ± 0.84 | 2.34 ± 0.97 | **0.045\*** | -0.44 | +0.19 | NA |
| Relative MP: Women (W/kg) | 20 | 1.76 ± 0.49 | 1.82 ± 0.67 | 0.57 | -0.18 | +0.06 | ns |
| Relative MP: Men (W/kg) | 16 | 2.50 ± 0.99 | 2.81 ± 1.02 | **0.025\*** | -0.74 | +0.31 | ns |
| Functional capacities (SPPB: x/12) | 35 | 9.23 ± 2.06 | 9.46 ± 2.18 | 0.32 | -0.22 | +0.23 | ns |
| Walking speed (m/s) | 36 | 0.92 ± 0.26 | 0.90 ± 0.27 | 0.42 | 0.17 | -0.02 | ns |
| Gait parameters (3-m TUG; s) | 37 | 13.8 ± 4.8 | 13.2 ± 4.3 | 0.06 | 0.37 | -0.67 | ns |

Results are presented as mean ± SD. Pre = before the 12-week intervention; Post = after the 12-week intervention; MP: muscle power; SPPB = short physical performance battery; TUG = timed up and go; 30-STs = 30 seconds sit-to-stand test; 5-STs = 5 repetitions sit-to-stand test; Muscle power = muscle power estimated via a validated equation; Relative muscle power = muscle power per body mass; delta change = post-pre; \*: *p*-value < 0.05; \*\*: *p*-value < 0.01; ES: effect size; ES is interpreted as follows: small (0.1-0.3), moderate (0.3-0.5) or large (>0.5); ns = no clinical change; + = clinical improvement; - = clinical deterioration; N/A = not applicable.

### Clinical changes following PACE intervention

Changes in clinical status regarding probable sarcopenia, fall risk and walking speed are summarized in **Table 2**, and **Figure 2**. No significant changes were observed in fall risk status or walking speed status between pre- and post-intervention (*p* > 0.05). In contrast, a significant shift in probable sarcopenia status was detected following the intervention, with a reduction in the proportion of probable sarcopenia participants and a corresponding increase in non-sarcopenic individuals (non-sarcopenic; pre: 50% vs. post: 72%, *p* = 0.011).

**Figure 2.**
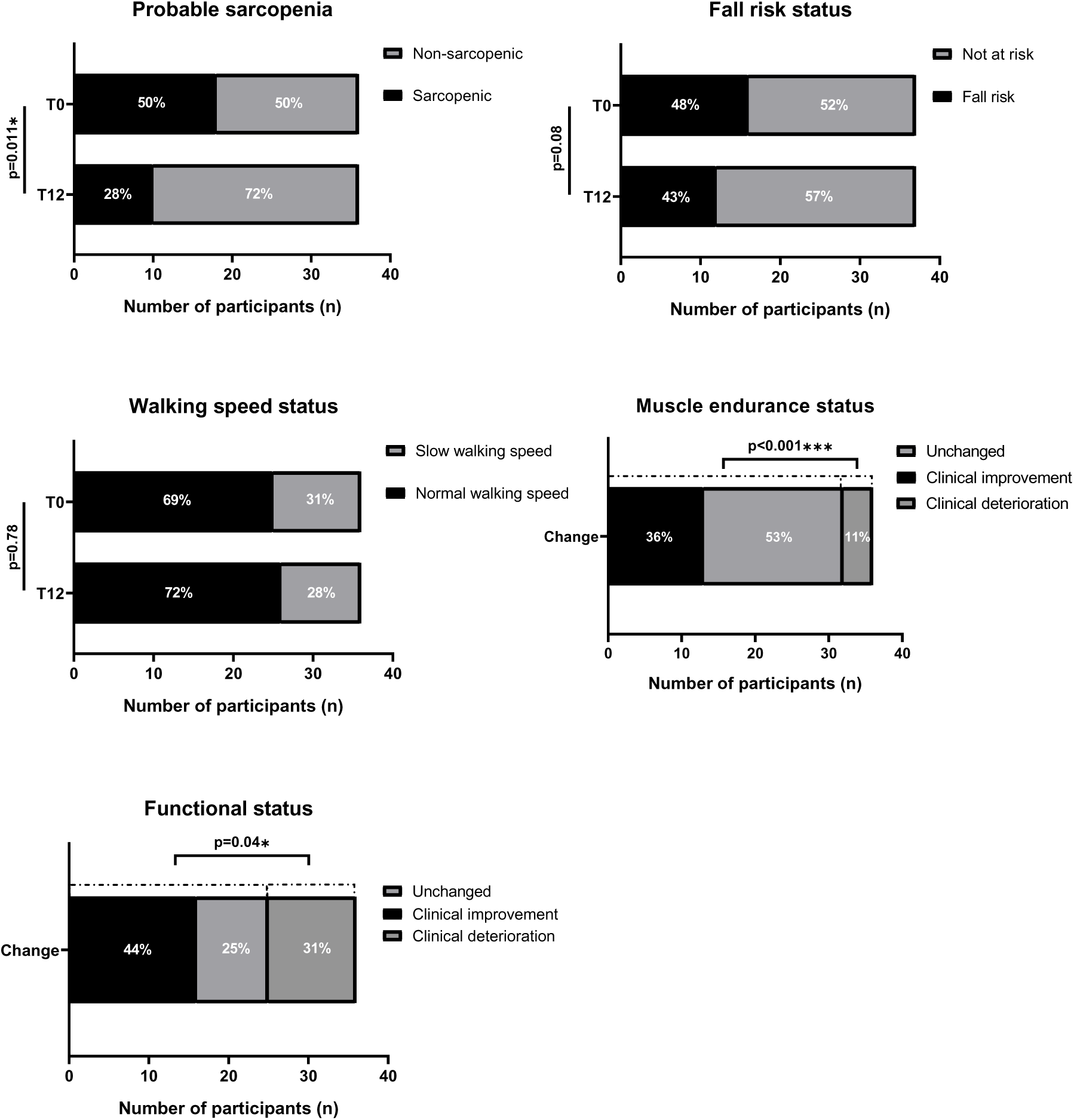
Clinical changes on physical performance outcomes in frail obese geriatric outpatients following a 12-weeks PACE program. Results are presented as counts and percentages. Sarcopenic: 5-rep STS **≥** 15 sec; Non-sarcopenic: 5-rep STS < 15. Fall risk: TUG **≥** 14; Not at risk: TUG < 14 sec; Normal walking speed: **≥** 0.8 m/s; Slow walking speed: < 0.8 m/s. Clinical deterioration for muscle endurance: 30-sec STS **≥** - 2points; Clinical improvement for muscle endurance: 30-sec STS **≥** + 2points. Clinical deterioration for functional status: SPPB **≥** - 1point; Clinical improvement for functional status: SPPB **≥** + 1point; SPPB = short physical performance battery; TUG = timed up and go; 30-sec STS = 30 seconds sit-to-stand test; 5-rep STS = 5 repetition sit-to-stand test.

**Table 2.** Clinical status changes and clinically meaningful changes following the 12-week PACE intervention in frail obese geriatric outpatients. *<u>Clinical status changes sub-part</u>*: Results presented as counts and percentages. Sarcopenic: 5-STS ≥ 15 sec; Non-sarcopenic: 5-STS < 15; Fall risk: TUG ≥ 14; Not at risk: TUG < 14 sec; Normal walking speed: ≥ 0.8 m/s; Slow walking speed: < 0.8 m/s; *: *p* < 0.05. McNemar test was used for statistical analysis. *<u>Clinically meaningful changes sub-part</u>*: Results are presented as counts and percentages. Improvement for muscle endurance: 30-STS ≥ + 2points; Deterioration for muscle endurance: 30-STS ≥ - 2points; Improvement for functional status: SPPB ≥ + 1point; Deterioration for functional status: SPPB ≥ - 1point; SPPB: short physical performance battery; 30-STS: 30 seconds sit-to-stand test; (+): Favorable (Improvement or Unchanged); (−): Unfavorable (Deterioration); *: *p* < 0.05; ***: *p* < 0.001. Exact binomial test was used for statistical analysis.

| CLINICAL STATUS CHANGES |  |  |  |  |
| --- | --- | --- | --- | --- |
| Status | Category | Baseline n (%) | Week 12 n (%) | *p*-value |
| Probable sarcopenia | Sarcopenic | 18 (50) | 10 (28) | **0.011\*** |
|  | Non-sarcopenic | 18 (50) | 26 (72) |  |
| Fall risk | Fall risk | 16 (48) | 12 (43) | 0.083 |
|  | Not at risk | 21 (52) | 25 (57) |  |
| Walking speed | Slow walking | 11 (31) | 10 (28) | 0.782 |
|  | Normal walking | 25 (69) | 26 (72) |  |
| CLINICALLY MEANINGFUL CHANGES |  |  |  |  |
| Status | Clinical change | n (%) | Difference | *p*-value |
| Muscle endurance | Improvement | 13 (36) | (+) vs. (-) | < **0.001\*\*\*** |
|  | Unchanged | 19 (53) |  |  |
|  | Deterioration | 4 (11) |  |  |
| Functional performance | Improvement | 15 (44) | (+) vs. (-) | **0.04\*** |
|  | Unchanged | 9 (25) |  |  |
|  | Deterioration | 11 (31) |  |  |

Analysis of clinically meaningful changes revealed that favorable outcomes, defined as either improvement or maintenance of performance, were significantly more frequent than deterioration for both muscle endurance and overall functional capacity. Specifically, 89% of participants achieved a favorable outcome in muscle endurance compared with 11% who deteriorated (*p* < 0.001). Likewise, favorable functional outcomes were observed in 69% of participants, significantly exceeding the proportion of those experiencing functional decline (31%; *p* = 0.04).

## DISCUSSION

The present study examined the effects of a 12-week unsupervised and adapted PA prescription in geriatric outpatients with overweight or obesity. Two main findings emerged: (i) participants exceeded current PA recommendations (∼244 min/week) without high drop-out rate (expecting: at least 25%) despite the absence of supervision, and (ii) clinically meaningful improvements were observed in lower-limb muscle function. Together, these findings confirm that the PACE tool is also feasible and effective for frail, overweight or obese older adults—a population traditionally considered difficult to engage physically ^25^.

Adherence to PA remains a major challenge in geriatric populations. According to the World Health Organization, adults should accumulate at least 150–300 minutes of moderate-intensity aerobic PA per week, or 75–150 minutes of vigorous-intensity activity, to achieve substantial health benefits ^31^. However, many older adults fail to meet these recommendations due to comorbidities, mobility limitations, fear of falling, low motivation, and limited access to supervised programs ^32^. In the present study, nearly two-thirds of participants (65%) accumulated more than 150 minutes of moderate physical activity per week. Given that achieving recommended PA levels is typically more challenging among individuals with overweight or obesity, this finding could be noteworthy. Interestingly, our participants reported a usability index (SUS) score of 75.3 ± 15.1%, indicating an acceptability of the PACE tool that aligns with the 70.8 ± 16.1% reported by Ruiz et al. ^25^. This finding is remarkable considering the marked differences in anthropometric status between the two cohorts.

Previous research indicates that obesity is associated with higher perceived exertion, impaired gait efficiency, reduced movement economy, and lower exercise tolerance ^33,34^. In addition, individuals with obesity tend to engage in less moderate-to-vigorous PA and more time in SB compared to their normal-weight counterparts ^35^ due to biomechanical and energetic constraints. In this context, the adherence observed in the present study suggests that tailored, clinically integrated pragmatic and adapted prescriptions may overcome barriers to PA participation in this population.

Our findings revealed also that the PACE program significantly improved lower-limb muscle function, as evidenced by gains in muscle function, (5-STS), muscle endurance (30-STS) and estimated muscle power. These improvements resulted to a significant reduction in the proportion of participants classified as probable sarcopenia. Probable sarcopenia is known to be a key determinant of disability, functional decline, and mortality in older populations ^36,37^. Accordingly, the observed shift from probable sarcopenic to non-sarcopenic status indicates that PACE produced not only statistically significant changes but also clinically meaningful benefits. The improvement in lower-limb muscle quality (function, endurance and power) is also known to be critical for preventing some age-related musculoskeletal declines ^38^, the activities of daily living and a strong predictor of functional capacities in older adults ^39,40^. Moreover, higher levels of lower-limb muscle function have been associated with a reduced risk of mortality, independently of age ^41^. Several physiological mechanisms may underlie these adaptations. Across the lifespan, individuals with obesity often display greater absolute maximal strength than their normal-weight peers ^42^. Thus, during exercises, carrying additional mass could increase self-loaded resistance and potentially amplifying adaptive responses to training by exposing the muscle lower limbs to greater mechanical tension and neuromuscular stimulation as proposed respectively by Thorén et al. ^43^ and Bosco et al. ^44^. One potential explanation about the functional gains observed is that majority of the PACE programs prescribed specifically feature lower-limb strength.

While the meta-analysis by Gómez-Redondo et al. (2024) supports the superiority of supervised exercise interventions for improving STS performance in older adults ^39^, the improvement observed in muscle endurance (30-STS: +1.17 reps) was comparable to the gains reported in several supervised interventions (Others (30-STS): +1.5 to +1.7 reps) ^45,46^. Our findings are also consistent with those of the MATCH study, which reported improvements in chair test performance following an unsupervised physical activity intervention in hospitalized older patients at risk of falls ^47^. Notably, the improvement in 30-STS performance was almost identical between studies (PACE: +1.17 vs. MATCH: +1.20 repetitions). The larger improvement in 5-STS observed in MATCH (MATCH: −5.77 vs. PACE: −1.18 sec), may be explained by the poorer baseline functional status of hospitalized older participants even if this finding (PACE: −1.18 sec) is closed to another supervised exercise trial (Other (5-STS): −1.02 sec) ^48^. In a pilot study (phase 1 of PACE co-creation), Ruiz et al. (2025) reported no significant clinical outcomes changes following a 12-week PACE program ^25^. The differences in functional gains observed between the two studies might be attributed to: 1) the specific characteristics of our cohort—which exclusively included overweight or obese geriatric outpatients; 2) the expansion of the SDT or ODT scores (from x/15 to x/18) that lead to differences in program intensity, and walking volume differences (brown program: ∼16% vs. Ruiz’study: ∼3%; level V: ∼35% vs. Ruiz’study: ∼3%) or/and; 3) the implementation of three variations per exercise that could lead to better adherence and execution of the prescription.

Furthermore, based on changes observed, the PACE tool seems also to be clinically relevant. Indeed, a substantial proportion of participants experienced favorable clinical trajectories in muscle endurance and functional status which are recognized to be associated with improved mobility and reduced risk of disability ^49,50^. The predominance of favorable outcomes in this cohort suggests that even an unsupervised, adapted PA prescription can elicit meaningful physical and functional adaptations in frail geriatric outpatient with overweight or obesity.

Overall, the present findings suggest that frailty and obesity should not be viewed as barriers to successful PA engagement. Instead, as previously shown ^25^, an adapted and pragmatic prescription model enables this sub-population to achieve recommended activity levels and derive meaningful physical benefits.

Several limitations should be considered when interpreting the present findings. As a pilot study, the sample size was relatively small, potentially reducing statistical power. Moreover, single-arm design without a control group limits the ability to attribute observed changes solely to the intervention and does not account for potential confounding factors or temporal effects. Thus, larger, adequately powered controlled trials including a control group are needed. Furthermore, the recruitment of participants from a single clinical setting may restrict the external validity of the results. Despite these limitations, the present study provides valuable preliminary data supporting the feasibility, acceptability, and potential effectiveness of the PACE intervention in older adults, thereby supporting the need for larger controlled trials to confirm these findings.

## CONCLUSION

The ability of PACE to elicit clinically meaningful improvements in muscle function as well as its implementability demonstrated that this pragmatic tool should be considered as an important geriatric routine care to counteract age-related sarcopenia among geriatric outpatients with frailty and obesity. Future research is warranted to establish the effectiveness of the intervention with or without obesity, explore long-term adherence, evaluate scalability within healthcare systems, and determine whether the observed benefits can be maintained beyond the intervention period.

## Author’s contributions

All authors read and approved the final version and agreed on the authorship order. **H.E.O:** Formal analysis, Data cleaning, Visualization, Writing - Original draft. **L.Y:** Data cleaning, Visualization, Writing - Review & Editing. **F.J.M.R:** Contribution to tool co-creation, Assessment, Investigation. **L.B:** Resources. **T.T:** Resources. **C.B:** Resources. **F.A.A:** Resources. **M.J.K:** Conceptualization, Tool co-creation, Methodology, Investigation, Visualization, Writing - Review & Editing. **M.A.L:** Conceptualization, Tool co-creation, Methodology, Investigation, Visualization, Writing - Review & Editing, Validation, and Supervision.

## Acknowledgements

We would like to express our gratitude to the patients, geriatricians, healthcare team, kinesiologists and research assistants who participated in this study and made possible to explore the integration of physical activity prescription into clinical practice.

## Data availability

The data that support the findings of this study are available from the corresponding author upon reasonable request.

## Supplementary material

**Supplemental Table 1.** Participants’ characteristics.

| Intervention | Completed | Drop-out | *P*-value |
| --- | --- | --- | --- |
| n (M/W) | 17/20 | 5/8 |  |
| Age (yrs) | 77.2 ± 5.7 | 75.0 ± 7.2 | 0.34 |
| BMI (kg.m<sup>-2</sup>) | 28.7 ± 3.2 | 29.9 ± 5.3 | 0.74 |
| ADL (/100) | 93.8 ± 12.9 | 93.1 ± 14.2 | 0.74 |
| IADL (/8) | 6.29 ± 2.12 | 6.54 ± 2.33 | 0.28 |
| MMSE (/30) | 22.8 ± 3.5 | 23.3 ± 6.9 | 0.76 |
| GDS-4 (/4) | 1.00 ± 1.32 | 0.85 ± 0.90 | 0.65 |

Data are presented as mean ± SD. n = number of participants; BMI: body mass index; ADL: activities of daily living; IADL: instrumental activities of daily living; MMSE: mini mental scale examination; GDS-4: geriatric depression scale.

**Supplemental Figure 1.**
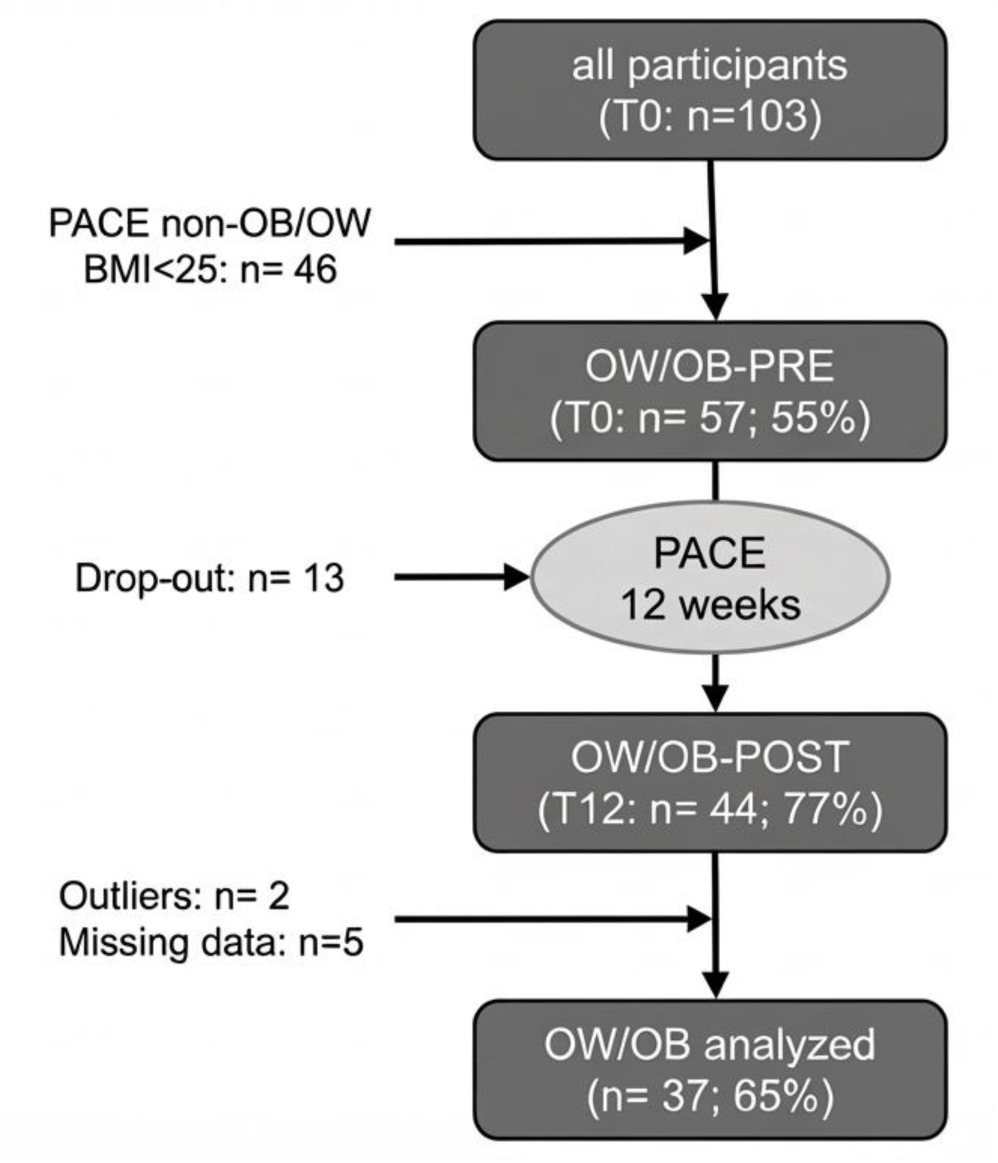
Study flowchart. OW=overweight; OB=obese; BMI=body mass index. PRE = before the 12-week intervention. Post = after the 12-week intervention. T0 = Baseline assessment. T12 = 12-week assessment.

**Supplemental Figure 2.**
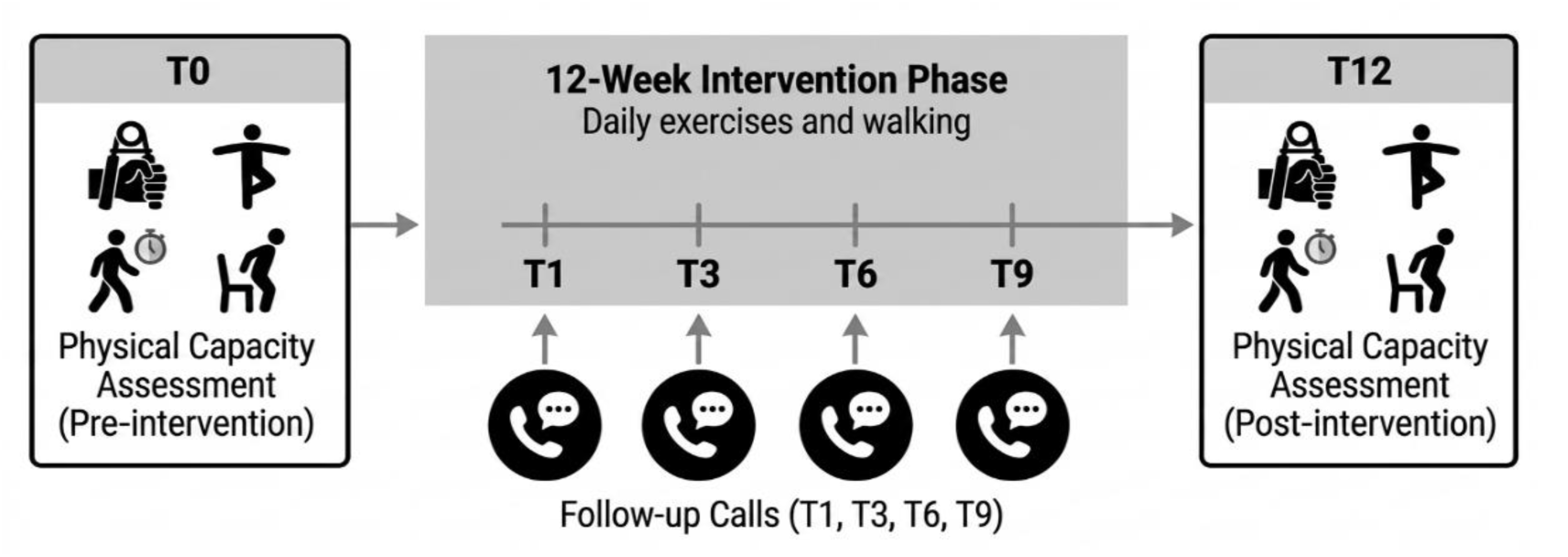
PACE Study Design. T0 = baseline assessment. T12 = 12-week assessment. Pre = before the 12-week intervention. Post = after the 12-week intervention. T1, T3, T6, T9 refer to follow-up calls at weeks 1, 3, 6, 9 respectively.

**Supplemental Figure 3.**
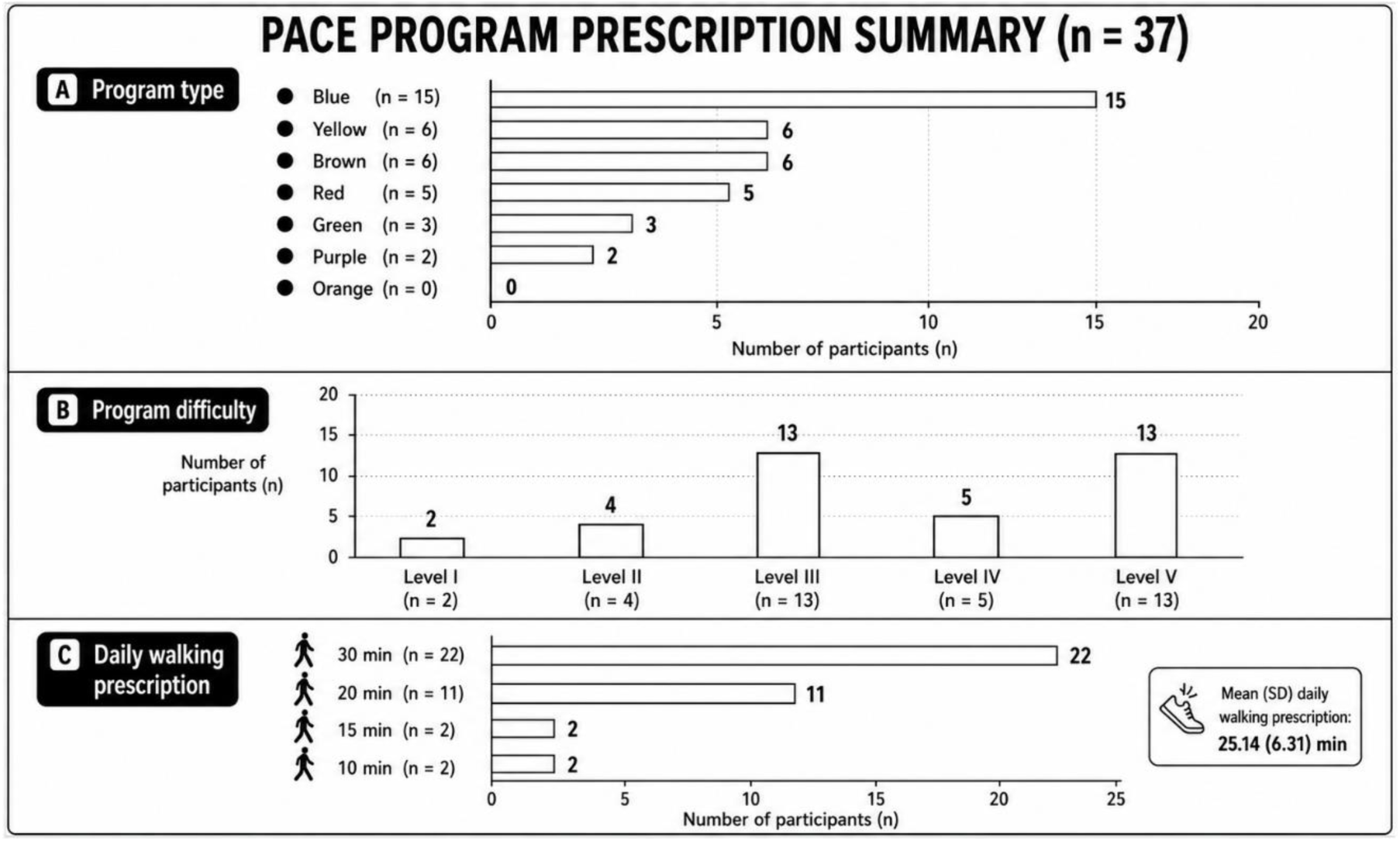
PACE program prescription. Blue = Strength; Yellow = Balance; Red = Flexibility; Green = Strength and Balance; Purple = Strength and Flexibility; Orange = Balance and Flexibility; Brown = Strength, Balance and Flexibility.

